# Female football specific energy availability questionnaire and menstrual cycle hormone monitoring

**DOI:** 10.1101/2021.10.29.21265667

**Authors:** Nicola Keay, Eddie Craghill, Gavin Francis

**Affiliations:** Department of Sport and Exercise Sciences, Durham University, United Kingdom; Women’s Team Support Doctor, Manchester United FC; Science4Peformance, London, United Kingdom

## Abstract

**Objectives:** The purpose of this study was to assess the energy availability status of professional female football players with an online Female Football Energy Availability Questionnaire (FFEAQ), combined with the clinical tool to model menstrual cycle hormones using artificial intelligence (AI) techniques.

**Methods:** The Female Football Energy Availability (FFEAQ) was developed based on published questionnaires, with a weighted scoring system to assess risk of Relative Energy Deficiency in Sport (RED-S). For menstrual cycle hormones AI techniques modelled hormone variation over a cycle, using the results from capillary blood samples taken at two time points.

**Results:** 21 female footballers of professional club level participated in this study, with mean age 22 years [range 16 to 30]. 20 athletes recorded positive scores on the FFEAQ, suggesting a low risk of Relative Energy Deficiency in Sport (RED-S). No players had experienced primary amenorrhoea. 5 athletes reported previous history of secondary amenorrhoea. Amongst the 15 players not taking hormonal contraception, 2 reported current oligomenorrhoea.

The application of AI techniques to model menstrual cycle hormones found that in 1 of the 3 players, subclinical hormone disruption was occurring with this player reporting variable flow of menstruation. Although the other 2 players showed expected menstrual hormone variation, 1 player reported variable flow according to training load, suggestive of subclinical anovulation. At the time of testing training load was low due to pandemic lock down.

**Conclusions:** The professional female football athletes in this study were found to be at low risk of RED-S from the FFEAQ. Modelling menstrual cycle hormones using AI techniques indicated that this has the potential to be an effective clinical tool in identifying subtle hormone dysfunction such as subclinical anovulatory cycles in female athletes.

**What are the new findings?:**

- Female football players can be at risk of low energy availability and development of the adverse health and performance consequences of Relative Energy Deficiency in Sport (RED-S)
- Sport specific screening questionnaires are a valuable clinical screening tool to identify those at risk of RED-S, to direct swift and personalised support to prevent progression from low energy availability to the clinical syndrome of RED-S
- Modelling menstrual cycle hormones with artificial intelligence (AI) techniques is an effective clinical tool to provide finer detail of hormone networks to identify subclinical hormone dysfunction in female athletes

**How might this study impact on clinical practice in the future?:**

- Female Football Energy Availability Questionnaire (FFEAQ) is a useful clinical screening tool to identify athletes at risk of RED-S
- Application of artificial intelligence to menstrual cycle hormones can provide a complete picture of hormone function. This clinical tool has the ability to detect subclinical hormone dysfunction as a precursor to developing functional hypothalamic amenorrhoea (FHA) in RED-S
- This AI clinical tool can also be helpful for athletes recovering from FHA to guide the appropriate return to full training once full hormone function is restored
- This AI hormone clinical tool can be used in distinguishing hypothalamic issues found in low energy availability; from reduced ovarian responsiveness found in perimenopause.

## Introduction

Endurance athletes are well documented to be at risk of low energy availability and hence the adverse health and performance consequences of Relative Energy Deficiency in Sport (RED-S)[1]. Nevertheless, emerging evidence and discussion of athletes participating in team sports, such as women’s football, suggests that they can also be at risk, whether low energy availability (LEA) arises intentionally or unintentionally, due to high training load and social pressures[2].

The validated Low Energy Availability Female Questionnaire (LEAF-Q)[3] focuses on the physical health aspects of low energy availability in female athletes. However, to date there are no sports specific screening questionnaires to identify female football players at risk of the health and performance consequences of RED-S.

A Sports Specific Energy Availability Questionnaire with clinical medical Interview (SEAQ-I) for male cyclists has been validated against bone mineral density (BMD) of the lumbar spine and hormones[4]. Furthermore, educational interventions in this research study proved effective in reversing the adverse health and performance consequences of those cyclists at risk of RED-S[5]. A Dance Energy Availability Questionnaire (DEAQ) for female and male dancers indicated that physiological drivers play a part in development of LEA and subsequent risk of RED-S[6].

Early identification of athletes at risk of LEA, would help in the prevention of RED-S. In females the endocrine network of the menstrual cycle is particularly sensitive to LEA. Whilst functional hypothalamic amenorrhoea (FHA) is the end point of LEA, more subtle dysfunction such as subclinical anovulatory cycles and luteal phase deficits may proceed this^7^. To date current menstrual cycle tracking may not detect these subtle endocrine disruptions.

The research objectives were to develop a female football energy availability questionnaire (FFEAQ) to identify players at risk of LEA and to test the clinical application of modelling female hormones over a menstrual cycle using artificial intelligence (AI) techniques.

## Methods

### Study design

A cross sectional study of professional female footballers at a UK football club from March to July 2021. All participants provided informed consent.

### Female Football specific health questionnaire

Participants were invited to complete a female football specific questionnaire (FFEAQ) which had been developed based on current clinical screening questionnaires.

### Blood biomarkers

Blood biomarkers were assessed for those athletes not on hormonal contraception and who were menstruating, using capillary blood samples, taken on day 14 and day 21 of a menstrual cycle. Samples were analysed for FSH (follicle stimulating hormone), LH (luteinising hormone), oestradiol and progesterone, at an accredited laboratory using a cobas8000 machine.

### Statistical analysis

Data analysis was performed using the open-source tools, SciPy and Pandas (NumFOCUS, Austin, Texas), implemented using the Python programming language. Summary statistics, including count, mean and standard deviation of responses, were calculated for the sample according to the responses to the questions. Body mass index (BMI) was calculated by dividing mass in kilogrammes by the square of height in metres. A minimum BMI (BMI min) was calculated based on the player’s minimum weight for current height. A weight variability value was calculated by dividing the difference between the maximum and minimum weights for the current height by the current weight.

A RED-S Risk Score was calculated by applying a points system to questionnaire responses particularly relevant to the syndrome (see RED-S Risk Score calculation supplementary file 2). Relevant factors included BMI, hormone levels, diet, injury history, measures of wellbeing (freshness, sleep, gut issues), attitudes to controlling diet and weight and any diagnosed eating disorder.

The modelling of the time variation of female hormones drew from the field of Artificial Intelligence. A tested system used Bayesian Inference to infer the best fit curves of the four key female hormones. The analysis was performed for each individual, taking into account reported cycle length and the measured hormones at two time points in the menstrual cycle. The output was used to assess overall hormone network function.

## Results

The questionnaire received complete responses from a total of 21 female footballers of professional club level, including 7 national team players.

The mean age of the players was 22 years [range 16 to 30]. On average, the players started playing football aged 6 and transitioned to full-time training at 18.

### Female Football Energy Availability Questionnaire (FFEAQ)

Anthropomorphic data are provided in Table 1: Anthropomorphic data. Weight variability is the range of minimum to maximum weight for current height divided by current weight. A value of 8% would be consistent with weight varying by +/-4%, so a player weighing 64.6 kg might have seen her weight vary between 62kg and 67kg and her BMI vary from 22.0 to 23.8.

**Table 1:** Anthropomorphic data.

|  | Height (m) | Weight (kg) | BMI (kg/m<sup>2</sup>) | Weight variability |
| --- | --- | --- | --- | --- |
| **Players (n=21)** | 1.68 [1.58 to 1.80] | 64.6 [57.0 to 77.0] | 22.8 [19.9 to 27.9] | 8% [3% to 25%] |

The sample included a range of playing positions shown in Table 2.

**Table 2:** Playing positions.

| Playing position | Forward | Midfield | Defender | Goalkeeper |
| --- | --- | --- | --- | --- |
| **Players** | 6 | 4 | 8 | 3 |

Most players had been actively engaged in sports prior to taking up full time football, typically training around 9 hours a week. Current weekly activity levels were consistent with professional athletes shown in Table 3

**Table 3:** Current weekly activity levels.

| Hours per week | Matches | Training | Gym | Total |
| --- | --- | --- | --- | --- |
| **Players** | 2.5 [0 to 5] | 12.9 [8 to 22] | 6.6 [2.5 to 13] | 22.0 [13 to 38] |

### Psychological factors

#### Attitudes to training

Eight respondents reported feeling worried about missing a session, while 13 noted that these things happen.

#### Attitudes to weight

Players reported on factors influencing attitudes to weight. Nine had been told to lose weight at some point in their training/professional career. 15 of the 21 players were not influenced by social media (e.g. Instagram) to try to lose weight, though 6 players were influenced to some extent.

Players weighed themselves on average 1.9 times a week. Half did not weight themselves at all, but one player weighed herself 20 times a week. Players’ self-esteem was generally not strongly affected by their ability to control what they eat and what they weigh, using a scale of 1 (no effect) to 6 (very important) shown in Table 4.

**Table 4:** Control and self-esteem.

| Impact on self-esteem | Controlling what you eat | Controlling what you weigh |
| --- | --- | --- |
| Mean score | 2.4 | 2.6 |

Players felt they were close to the best weight, with no desire to be significantly lighter or heavier.

### Menstrual History

Female hormone history was investigated in some detail. No players reported primary amenorrhoea: all players had experienced the commencement of menstruation, though this was not until after the age of 15 in four of the players. Secondary amenorrhoea: menstruation had ceased for six months or more, at some point in the past, for five players. Oligomenorrhea: of the 15 players not using hormonal contraceptives, 2 experienced fewer than nine cycles per year.

All menstruating players felt that their cycles affected their performance, primarily during the menstrual bleed (13) or in the two weeks before or just before. Bleed length was either 3-4 days (8) or 5-7 days (7). Eleven players used menstrual tracking apps.

On being asked if the hormones in the oral contraceptive pill are equivalent to their own hormones, 7 said no, 12 did not know and 2 said yes. Of the six players, using hormonal contraceptives, their reasons were contraception, reduction of menstrual period pain or to regulate cycles.

There were mixed opinions on whether it was normal for female athletes not to have periods: 12 said no, 5 yes and 4 did not know. Twelve players associated negatives with not having periods, 7 did not and 2 did not know.

### Illness and injuries

Most players had taken time off due to injury in the last year. Injuries tended to be soft tissue related, though 6 players had fractured bones. Most time loss to injury had been 14 days or less, though some rehabilitated for much longer, these tending to be associated with fractures. Players had had relatively little illness: on average just over 2 off days in the year.

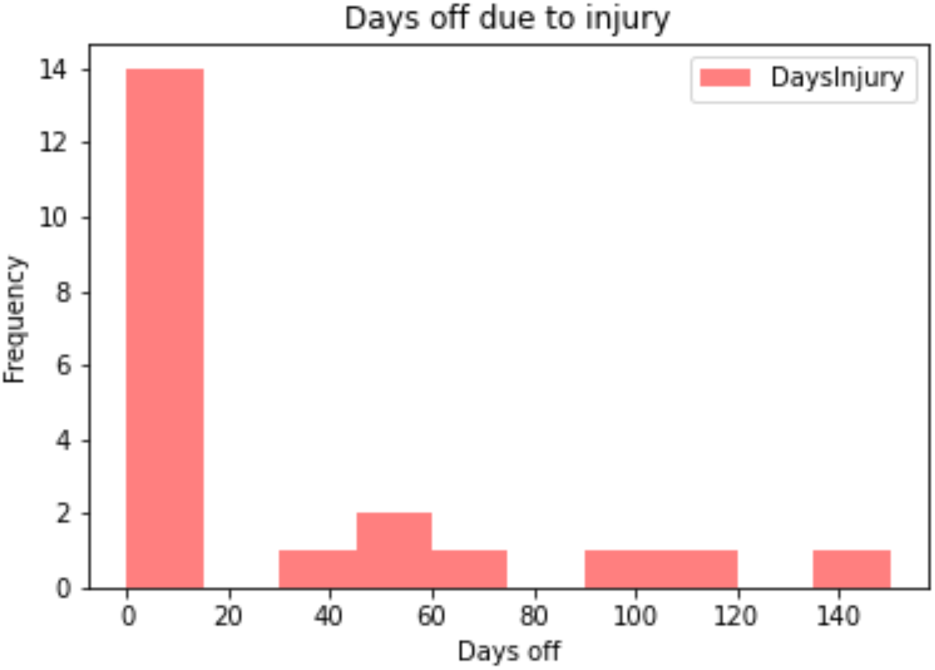

### Medications, supplements, allergies/intolerances

Players were generally not using medications or supplements. Four players reported minor allergies.

### Eating habits

Average caffeine and dairy consumption were generally low: one cup of coffee and three portions of dairy. Two players were vegetarian, but none were vegan. Other than one player who avoided fish and one who limited carbohydrates, the large majority of players (17) did not exclude food types. Players benefited from advice provided by the team/club access to a dietician/nutritionist.

### Smoking

None of the players smoked.

### Wellbeing

On a rating scale for freshness from 1 (extremely fatigued) to 6 (no fatigue at all), players scored 3.2, on average. Using a similar scale for sleep from 1 (hardly ever get a good night’s sleep) to 6 (always), players ranked 3.6 on average. Among those having problems sleeping, common reasons were difficulty falling asleep (8), early waking (7) and disrupted sleep (3).

Ratings for the digestive system from 1 (continuous problems) to 6 (none) produced an average of 4.0. 11 reported no digestive problems, while 7 had bloating, 2 suffered discomfort and 3 constipation.

### Eating disorders

Only one player had a previous diagnosis of an eating disorder. There was a range of awareness of medical terms.

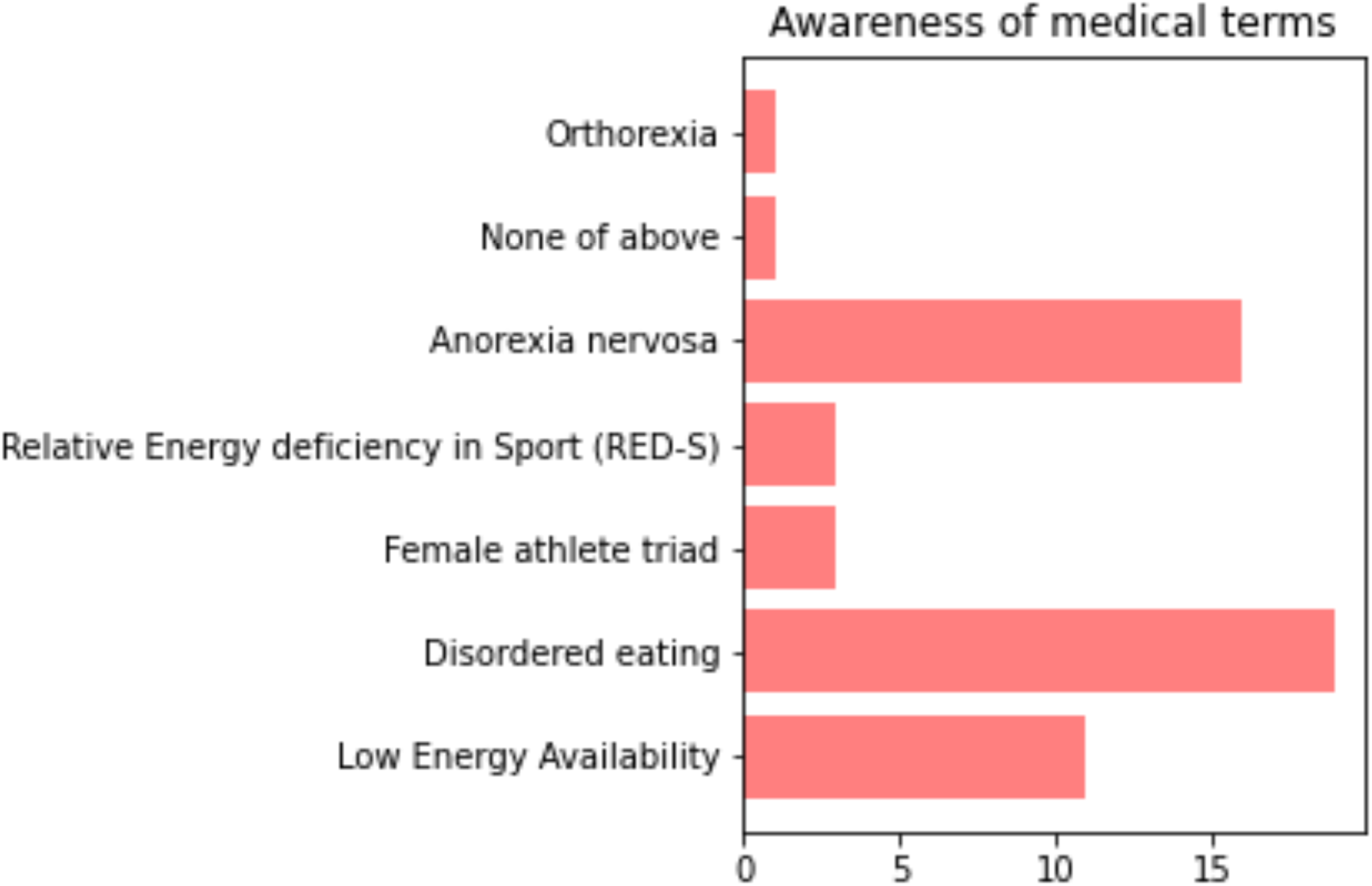

RED-S Risk Scores ranged from -2 to +12, with negative scores indicating a higher risk. The average score was +6.4, indicative of healthy status. The only player with a negative score had experienced some menstrual disruption and injuries, as well as strong desires to control her food and weight. Nevertheless, her current BMI was healthy, and she was not experiencing problems with fatigue, sleep or gut issues.

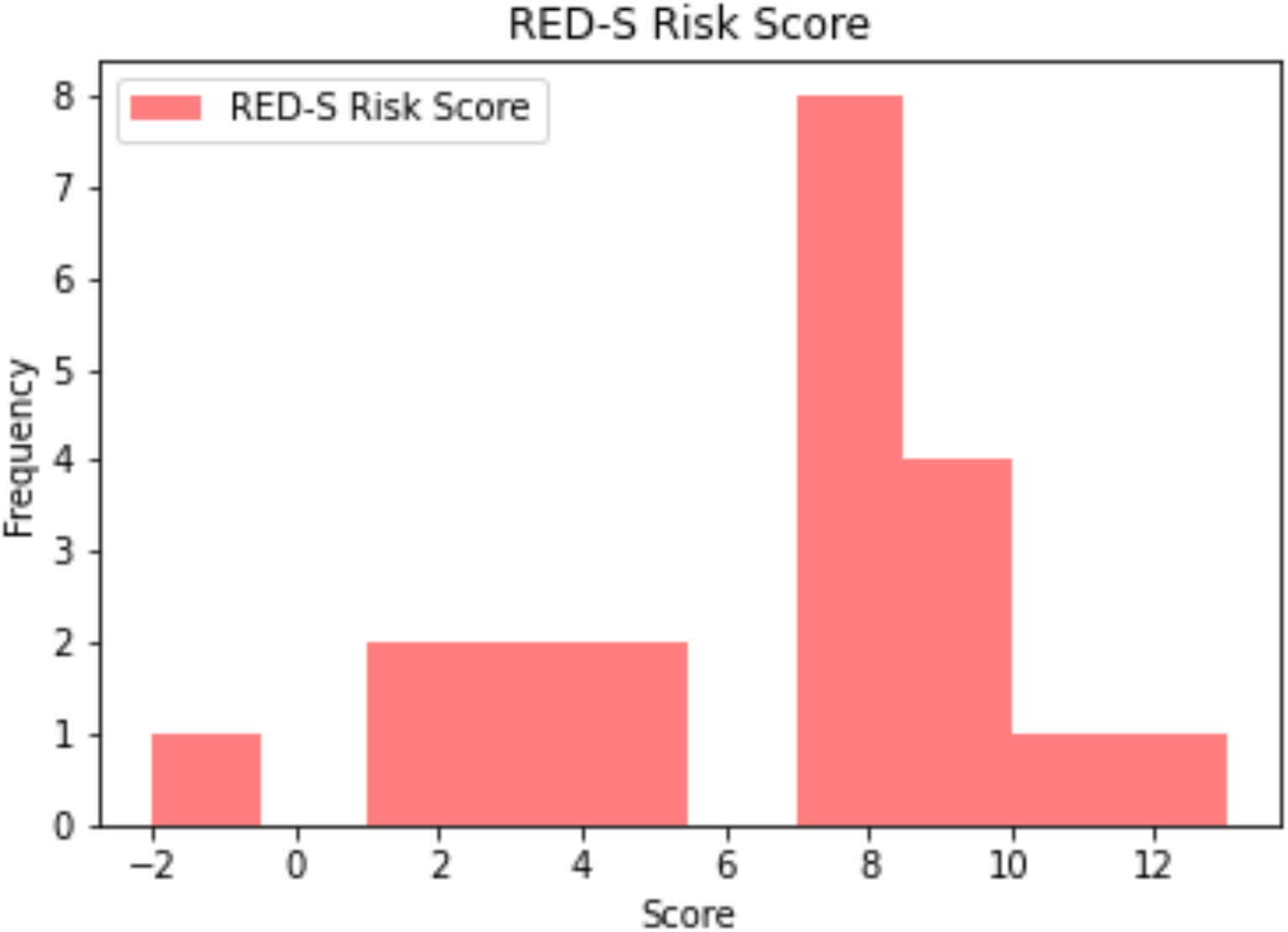

### Female Hormone Modelling

3 players performed female hormone modelling. For 1 player, subclinical hormone disruption was occurring demonstrated by the modelled time course of hormone fluctuations over the entire menstrual cycle for FSH, LH, oestradiol and progesterone, in keeping with subclinical anovulation. Furthermore, this player reporting variable flow of menstruation which could be clinical consequence of low range oestradiol over the cycle. Although the other 2 players showed expected menstrual hormone variation, 1 player reported light flow according to training load, suggestive of subclinical anovulation. At the time of collecting sample, training load was unusually low due to pandemic early lock down.

## Discussion

This study found that female football players can be at risk of low energy availability, which can be identified with a female football specific questionnaire (FFEAQ). Artificial intelligence techniques can be used to model the hormones of the menstrual cycle from two capillary blood samples taken during the cycle which can detect early subclinical emergent female hormone network dysfunction.

### Female Football Energy Availability Questionnaire (FFEAQ)

The clinical assessment of energy availability in female footballers has historically been overlooked and under-utilised. In an elite environment, individual indicators of energy availability are routinely detected via the multi-disciplinary team however the bigger picture can remain challenging to piece together. The fewer the resources available to the medical team, the greater the challenge.

Diagnostic accuracy can also be compromised by the use of hormonal contraceptives such as the combined oral contraceptive pill (COCP) due to its effect on down regulation of the hypothalamic-pituitary-ovarian axis and consequent “switching off” of menstruation as a barometer of internal health female hormone network function. Even where there is low energy availability, the exogeneous female hormones in the COCP will induce a regular withdrawal bleed when taken as routinely prescribed. This can erroneously be regarded as a marker of a functioning hypothalamic system and mask the extent of RED-S[8].

The FFEAQ has the advantage of being a comprehensive clinical tool, which takes into account other indicators of low energy availability, beyond menstrual function and furthermore, includes sports specific factors. Attempts to evaluate energy availability in other sports, primarily those more traditionally associated with RED-S, have produced the female athlete screening questionnaires for low energy availability: LEAF-Q[3]. However, the LEAF-Q does not consider football specific factors, such as playing position, nor diet, injury type, wellbeing metrics or psychological factors. Furthermore, some of these factors are included in the FFEAQ weighted scoring system.

Other questions in the FFEAQ can be used for sub analysis in a longitudinal study of professional players in terms of playing position and breakdown of training load. The evidence is not currently robust enough to indicate that defenders are more at risk of RED-S than forwards for instance. There is however evidence to suggest the physical load on goalkeepers is less than outfielders, so correcting the FFEAQ score by 0 for an outfielder and +1 for a goalkeeper is proposed. This adjustment has the potential to provide further detail in the risk assessment of low energy availability in female football players.

The FFEA-Q standardises the assessment of health and energy availability in a way which can be used by medical practitioners and female footballers themselves at all levels of the game. The weighted scoring system of sports specific questionnaires also allows direct comparison between female athletes who train in other sports[6]. This provides finer detail, compared to the more generic LEAF-Q.

Using a threshold of -1 or less for “high risk of RED-S”, only one athlete scored within this category. This is consistent with the low incidence of low energy availability expected in women’s football when compared to some other sports. It is also indicative of an environment and a culture where steps had already been taken to address this issue.

A further five participants scored 4 or less. When correlated to prior multi-disciplinary assessment of energy availability, all but one had previously been identified as at risk by other means. This suggests the FFEA-Q is at least as effective at predicting those at risk compared to standard methods. This may be because certain psychological or behavioural factors are more challenging to elicit in face-to-face discussion. These findings support the creation of an “at risk of RED-S” category for those scoring 0-4.

Recognition is required that the FFEA-Q provides an evaluation of energy availability only at the time it is administered. Whilst respondent’s beliefs and previous experiences are accounted for, they are weighted sufficiently to allow normalisation of scores should subsequent behaviours and beliefs alter. This is evidenced by healthy scores amongst two participants who were previously given a diagnosis of RED-S associated with bony injuries but received subsequent MDT input to correct this.

Ultimately, clinical assessment will remain the gold-standard for the assessment of energy availability in female footballers however this can be enhanced through use of the FFEA-Q and the novel usage of individualised hormone profiling.

### Artificial Intelligence in modelling female hormones

Tracking the menstrual cycle, is a good starting point as an indirect indicator of female hormone health. However, subclinical anovulatory cycles can be missed. This is of concern as this situation, if left undetected, can have adverse health consequences[9]. Whilst blood testing is the “gold standard” for assessing the concentration of all four of the key female hormones, individual differences in hormone timing, means that subtilties of dysfunction can be missed[^10^]

Application of artificial intelligence techniques to biological systems and specifically in clinical medicine makes this a valuable clinical tool which is being used to personalise health care and medicine[11]. This approach is clinically effective for early identification of hormone disruption due to imbalances in training load and nutrition in professional dancers[12]. This clinical tool can also be used to guide return to training after restoration of energy availability. Quantification of dynamic female hormone function can also be valuable in monotiling the effect of training load over a season. Furthermore, this approach can be used to distinguish hypothalamic down regulation from reduced ovarian responsiveness in older age group athletes.

### Limitations and further work

Although all the athletes came from the same professional team, self-selection bias could play a part. Further work will be to undertake a longitudinal study of players to assess the impact of a full season on FFEAQ scores and menstrual cycle hormones.

The AI based clinical tool of modelling menstrual cycle hormones could also be applied in the personal monitoring of the effective restoration of endocrine networks following FHA due to RED-S to guide return to play. For retired female football players, based on symptoms and a static, single blood test, it is challenging to make the important clinical distinction between hypothalamic down regulation declining ovarian response occurring during perimenopause in lead up to menopause. AI assisted modelling of female hormone networks has the potential to be clinical tool to distinguish between the effects of low energy availability and physiological change on endocrine networks.

## Conclusions

A sport specific, female football energy availability questionnaire (FFEAQ) is a useful screening tool, which can be used, combined with clinical input to identify, and support athletes at risk of low energy availability and developing RED-S. The application of AI techniques to model female hormone networks can personalise clinical advice.

## Data Availability

All data produced in the present study are available upon reasonable request to the authors

## Statements

## Acknowledgments

Thank you to the athletes, staff and the club for their interest and participation in this study.

## Contributors

NK EC and GF: conceptualisation of project, development of study design, involvement of athletes, drafting and revision of manuscript. GF: advanced statistical analysis drafting and revision of manuscript.

## Ethical Approval

This study was reviewed and approved by Durham University research ethics committee.

## Female Football Energy Availability Questionnaire. Supplementary file 1

This online questionnaire should take about 15 minutes to complete. The purpose of this project is to optimise the health and performance of athletes so that they are able to sustain long, successful careers. Currently there are no health screening questionnaires specific to type of sport. This study proposes to assess health in female footballers.

Project title: Assessment of Health and Performance in Athletes. Researchers: Dr Nicky Keay (medical doctor), Dr Eddie Craghill (medical doctor), Gavin Francis (data analyst). Ethical approval from Durham University 27/02/20

### Consent

In order to complete this questionnaire, we request that you confirm that you understand the purposes of the project, what is involved and that you are happy to take part. Please click on this link for the Information Sheet and Privacy Notice. I confirm that I have read and understand the online Information Sheet and the online Privacy Notice for this project. I have had sufficient time to consider the information and ask any questions I might have and I am satisfied with the answers I have been given I understand who will have access to personal data provided, how the data will be stored and what will happen to the data at the end of the project. I understand that anonymised versions of my data may be archived and shared for legitimate research and educational purposes. I understand that my participation is entirely voluntary. I confirm that I am happy to take part in this project by answering this online questionnaire.

### Football Background (1 of 7)

Surname

First name

What is your date of birth?

Please select your favoured playing positions

> Goalkeeper
>
> Central defender
>
> Full/Wing back
>
> Central midfielder
>
> \Winger
>
> Striker
>
> Other…

At what level do you play on regular basis?

> Domestic team
>
> National team

Age you started playing football?

Age you transitioned into full-time football programme?

Please indicate your football activity in hours per week 0-5 6-10 11-15 15-20 20-25 26+

> Training
>
> Matches
>
> Conditioning

Before full-time football training, did you take part in any sports? If so please state which sport(s) and number of hours per week.

> Weight bearing exercise
>
> Resistance exercise
>
> Non weight bearing exercise

How do you feel if you have to miss training?

> Relieved, I need more rest
>
> These things happen
>
> Worried/anxious

### Medical History (2 of 7)

Current height (in metres, e.g. 1.65)

Current weight (kg)

Lowest weight for current height (kg)

Highest weight for current height (kg)

How often do you weigh yourself per week away from the club?

### Female hormones (3 of 7)

Age your periods started?

> 13 years or younger
>
> 14-15 years
>
> 16 years or older
>
> Never started

Have your periods stopped for 6 or more consecutive months (besides pregnancy or taking hormonal contraception)?

> No, never
>
> Yes, it has happened before
>
> Yes, that is the situation now

If your periods stopped at some time in your life, for how many months did this last? Please indicated if you have been diagnosed with any of the following?

> Polycystic Ovary Syndrome (PCOS)
>
> Endometriosis
>
> Polycystic Ovary Syndrome (PCOS)
>
> Endometriosis

Do you think the hormones in the oral contraceptive pill are equivalent to your own hormones?

> Yes
>
> No
>
> Don’t know

Have you ever been pregnant?

> Yes
>
> No

Are you currently taking any form of hormonal contraceptive?

Yes

No

### Hormonal contraception

If yes to taking hormonal contraception. Then go to section 4 Medications and lifestyle

What type of hormonal contraception are you using?

What is the reason you are taking hormonal contraception?

> Contraception
>
> Reduce menstrual period pain
>
> Decrease menstrual period bleeding
>
> To regulate cycle
>
> Otherwise no bleeding occurs

### Menstrual Cycles

If no to taking hormonal contraception

Please indicate the frequency of your cycles over the last year

> 9 or more periods at regular intervals (eg regularly at 25 to 34 day intervals)
>
> 9 or more periods at irregular intervals (sometimes a week early or late)
>
> Fewer than 9 periods
>
> No periods for 6 months

For you, what is the average numbers of days between one menstrual cycle and the next? Do you feel your menstrual cycles affect your physical performance and/or wellbeing?

> No impact
>
> In the two weeks before the menstrual bleed
>
> During the menstrual bleed
>
> Mid cycle

What best describes the average length of your menstrual bleed?

> 1-2 days
>
> 3-4 days
>
> 5-6 days
>
> 7-8 days
>
> more than 8 days

What best describes the nature of your menstrual flow?

> Generally light (requiring light sanitary wear)
>
> Generally heavy (requiring maximum sanitary wear)
>
> Neither light, nor heavy
>
> Very variable

Does a high training load change the nature of your menstrual cycle?

> Does not affect my cycle
>
> Lighter bleed for fewer days
>
> Heavier bleed for more days
>
> Menstruation stops

If you use a mobile application to track your periods, which app do you use?

### Medications and lifestyle (4 of 7)

Please list any prescribed medications

Please list below any allergies or food intolerances

What is your average caffeine intake per day? (Equivalent cups of caffeinated coffee) How many average portions of dairy do you consume per day. Portion = milk on cereal/tea/coffee, cheese, yoghurt)?

Are you vegetarian?

Are you vegan?

Do you exclude any types of foods by choice?

> Carbohydrates
>
> Fats
>
> Meat
>
> Fish
>
> Dairy
>
> Gluten
>
> Other

Have you been told by coaches/nutritionists to exclude any foods? If so, please list and provide reasons, if known.

Do you smoke?

Have you ever smoked

### Injuries (5 of 7)

During the last year how many soft tissue injuries, e.g. muscle, ligament, tendon, joint (excluding fractures) have you had?

Of these soft tissue injuries, how many were recurrent (i.e. in the same place, or same type of injury)?

How many bone injuries have you had since playing football full time? This includes bone stress response injuries, stress fracture and full break fracture.

If you had bone injuries how many of these were recurrent, i.e. same place, or same type of injury)?

If you have had any type of fractures, where have these been located?

> Legs
>
> Feet
>
> Pelvis
>
> Spine
>
> Arms

If you experienced any type of injury, did your treating healthcare professional ask you about your periods?

### General Health (6 of 7)

During the last year how many days off football have you had due to illness? How would you rate your levels of freshness over the past year?

> Score 1-5

How would you rate your sleep quality over the past year?

> Score 1-5

What type of sleeping difficulties do you experience?

How would you rate your digestive system over the past year?

What type of digestive problems do you experience (not related to menstruation)?

> None
>
> Constipation
>
> Bloating
>
> Discomfort

How often do you have bowel movements, on average?

> Several times a day
>
> Once a day
>
> A few times per week. Less frequently than daily
>
> Once a week or more rarely

How would you describe your normal stool?

> Normal (soft)
>
> Like diarrhoea (watery)
>
> Hard and dry

### Nutrition (7 of 7)

Which sources do you consult for advice on nutrition?

> None
>
> Internet
>
> Friends/coaches at my team/club
>
> Team/club provided access to dietician/nutritionist
>
> Professional dietician/nutritionist

Have you been told to lose weight at any point in your training/professional career?

Do social media (eg Instagram) make you feel you should try to lose weight?

How does controlling what you eat/what you weigh affect the way you feel about yourself? At what weight (in kg) do you play your best football?

Do you think you are more likely to be in starting line up if you are low weight/fat for your height?

Please indicate which of these terms you have heard of

> Low Energy Availability
>
> Female athlete triad
>
> Relative Energy deficiency in Sport (RED-S)
>
> Disordered eating
>
> Orthorexia

Have you ever been diagnosed with an eating disorder?

Do you think it is normal for female athletes not to have periods?

Apart from not being able to get pregnant, do you think there are there any negatives to women not having periods?

### RED-S Risk Score calculation. Supplementary file 2

The RED-S Risk Score was calculated as the sum of positive and negative points assigned to responses, as show below.

BMI +1 unless <20 -1 or <18 -2

BMI min +1 unless <20 -1 or <18 -2

How often do you weight yourself per week? >6 -1

First period ‘11 years or younger’: +1, ‘12-13 years’: +1, ‘13-14 years’: +1, ‘15 years or older’: -1, ‘Never started’: -2

Are your menstrual cycles regular? ‘Yes (9 or more per calendar year)’: +1, ‘No (less than 9 per calendar year)’: -1

Have your periods stopped for 3 or more consecutive months (besides pregnancy or taking hormonal contraception)? ‘No, never’: +1, ‘Yes, it has happened before’: -1, ‘Yes, that is the situation now’: -1

Are you vegetarian? ‘No’: +1, ‘Yes’: -1

Are you vegan? ‘No’: +1, ‘Yes’: -2

Carbohydrates (excluded from diet): -1

Do you smoke? ‘No’: +1, ‘Yes’: -1

During the last year how many days off training have you had due to injury? 0 unless >=7 days -1 or >=14 days -2

During the last year how many soft tissue injuries, e.g. muscle, ligament, tendon, joint (excluding fractures) have you had? 0 unless one injury -1 or more -2

Of these soft tissue injuries, how many were recurrent (i.e. in the same place, or same type of injury)? if >2 then -1

If you had bone injuries how many of these were recurrent, i.e. same place, or same type of injury? if >2 then -1

Fracture Legs -1

Fracture Feet -1

Fracture Pelvis -2

Fracture Spine -2

Fracture Arms -1

Extremely fatigued all the time vs No fatigue at all (scale 1 to 6) ‘1’: -2, ‘2’: -1, ‘3’: 0, ‘4’: 0, ‘5’: +1, ‘6’: +2

Hardly ever a good night’s sleep vs Always a good night’s sleep (scale 1 to 6) ‘1’: -2, ‘2’: -1, ‘3’: 0, ‘4’: 0, ‘5’: +1, ‘6’: +2

Continuous problems vs No problems (scale 1 to 6) ‘1’: -2, ‘2’: -1, ‘3’: 0, ‘4’: 0, ‘5’: +1, ‘6’: +2

Controlling what you eat (scale 1 to 6) ‘1’: +2, ‘2’: +1, ‘3’: 0, ‘4’: 0, ‘5’: -1, ‘6’: -2

Controlling what you weigh (scale 1 to 6) ‘1’: +2, ‘2’: +1, ‘3’: 0, ‘4’: 0, ‘5’: -1, ‘6’: -2

Have you ever been diagnosed with an eating disorder? ‘No’: +1, ‘Yes’: -2

